# Clonality and timing of relapsing colorectal cancer metastasis revealed through whole-genome single-cell sequencing

**DOI:** 10.1101/2022.03.03.22271700

**Authors:** Joao M. Alves, Sonia Prado-López, Laura Tomás, Monica Valecha, Nuria Estévez-Gómez, Pilar Alvariño, Dominik Geissel, Dominik Paul Modest, Igor M. Sauer, Johann Pratschke, Nathanael Raschzok, Christine Sers, Soulafa Mamlouk, David Posada

## Abstract

**BACKGROUND:** Recurrence of tumor cells following local and systemic therapy is a significant hurdle in cancer. Most patients with metastatic colorectal cancer (mCRC) will relapse, despite resection of the metastatic lesions. A better understanding of the evolutionary history of recurrent lesions is thus required to identify the spatial and temporal patterns of metastatic progression and expose the genetic determinants of therapeutic resistance.

**METHODS:** Utilizing a robust Bayesian phylogenetic approach, we analyzed a unique single-cell whole-genome sequencing dataset comprising 60 cells sampled from metastatic and recurrent hepatic lesions of a patient with a long-term disease course to investigate the origin, timing, and clonality of a colorectal metastatic relapse. We further tracked the changes in the size of the malignant cell population and evaluated the impact of the treatment strategy on the mutational landscape of this tumor.

**RESULTS:** Our results suggest that the recurrent lesion originated from the clonal expansion of a single drug-resistant metastatic lineage, which began to expand around one year before surgical resection of the relapse. We additionally observed substantial variability in the substitution rates along the tumor cell phylogeny and found a large number of mutations specific to the ancestral lineage that gave rise to the relapse, including non-silent mutations in CRC genes. Moreover, our results point to a substantial contribution of chemotherapy exposure to the overall mutational burden.

**CONCLUSIONS:** Our study suggests that resistant colorectal metastatic clones can quickly grow, even under strong drug-imposed pressure, highlighting the importance of profiling the genomic landscape of tumor lesions to identify mutations potentially contributing to treatment failure.

## Introduction

Metastatic colorectal cancer (mCRC) remains one of the most common causes of cancer death (Xi and Xu, 2021). While population-wide screening programs have led to significant improvements in the detection and treatment of early lesions (Levin *et al*., 2018), a large proportion of colorectal cancer patients present with distant metastatic disease at the time of diagnosis or after a disease-free period. In mCRC, the liver is the most frequent location of metastasis (Sorbye, 2014; van der Geest *et al*., 2015). At this stage, resection of liver metastases can improve survival (Mitry *et al*., 2008; Kopetz *et al*., 2009; Van Cutsem *et al*., 2016). However, 50-75% of patients still suffer a relapse of the disease, primarily as a local recurrence in the liver (D’Angelica *et al*., 2011; Imai *et al*., 2016).

Studies on the origin and mechanisms of metastatic recurrence have focused on blood and bone marrow cancers, where acquiring longitudinal samples throughout a patient’s disease is entirely feasible (Shlush *et al*., 2017; Sentís *et al*., 2020; Waanders *et al*., 2020). Studies of recurrence in solid tumors are less common due to the difficulty in obtaining longitudinal samples. Still, examples exist in non-small cell lung carcinoma (J. Chen *et al*., 2022), pancreatic ductal adenocarcinoma following surgery (Sakamoto *et al*., 2020), hepatocellular carcinomas (Ding *et al*. 2019), head and neck tumors (Roh *et al*., 2018), triple-negative breast cancer ((Kim *et al*., 2018)) and melanoma (Liu *et al*., 2021). The patterns of relapse vary between cancer types; however, most studies found recurrence after treatment to predominantly originate from clones already present in the pre-treated tumor. Despite numerous studies on the evolutionary relationship between primary CRC tumors and spatially or temporally distinct metastases (Leung *et al*., 2017; Ishaque *et al*., 2018; Alves *et al*., 2019; Hu *et al*., 2019; Nikbakht *et al*., 2020; H.-N. Chen *et al*., 2022), information on the subclonal pattern of recurring metastasis post-treatment in mCRC is lacking, and treatment continues to depend on routine clinical procedures.

Survival of tumor cells after surgical resection and therapy is a significant obstacle in treating most cancers. Understanding the evolution of recurrent lesions is critical to identifying the origin of resistant cells and the mechanisms behind cancer relapse, consequently helping to shape treatment strategies. This study investigates the evolution and potential determinants of metastatic relapse in mCRC by leveraging a unique single-cell dataset comprising whole-genome information from 60 cells collected from a liver metastasis and a recurrent liver lesion. Using a powerful phylogenetic approach, we were able to reconstruct the genomic history of the relapse, including its origins, age, and clonality, and identified the putative evolutionary and molecular processes behind the reestablishment of the tumor.

## Material & Methods

### Patient history and sample collection

In November 2008, a male in his 40s diagnosed with a rectal adenocarcinoma with synchronous liver metastasis (pT4N0M1) underwent resection of the primary tumor, followed by a liver resection in December 2008 for removal of his liver metastases. From February 2009, the patient was treated with eight cycles of adjuvant chemotherapy with capecitabine and oxaliplatin. In February 2010, a PET-CT scan revealed a new lesion in the liver, with histological assessment confirming a metastatic adenocarcinoma of colorectal origin. After that, seven cycles of FOLFIRI and Bevacizumab (VEGF inhibitor) were administered, from March to June 2010, leading to stabilization of the disease. In February 2011, two years following the first liver resection, the recurrent liver lesion was surgically resected. An overview of the patient’s clinical course during this period is depicted in **Fig. 1a**. The disease was further complicated by lung metastases, treated surgically in 2014, a local recurrence in 2015, and recurrent liver, lung, and bone metastases in 2018, which were treated with palliative radiation until the patient passed away in 2020.

**Figure 1.**
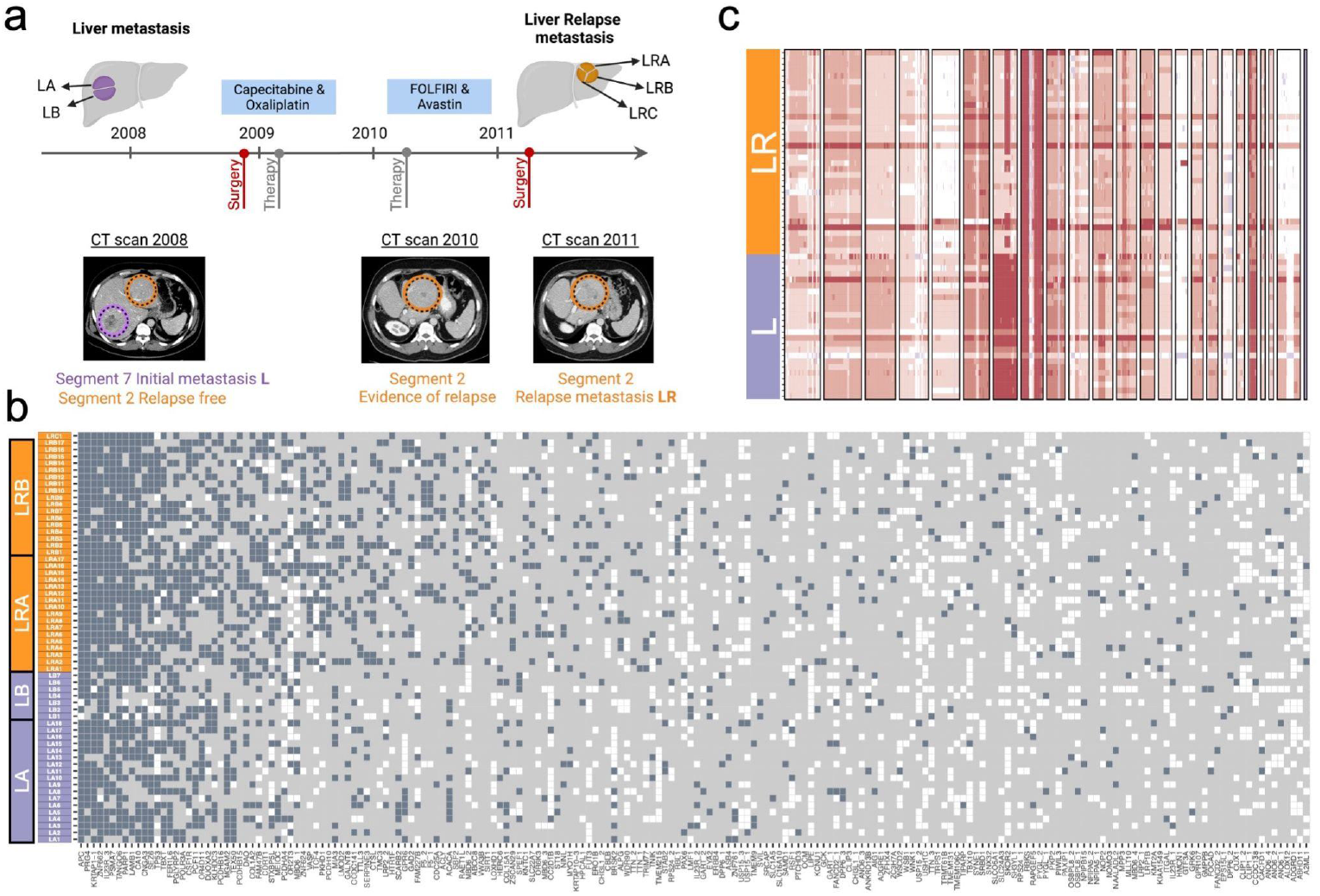
Overview of clinical progression and genomic characterization of single-cell data obtained from longitudinal metastatic biopsies. **a**. Clinical course of disease with the timeline depicting the year at which the patient underwent surgical resection - 2008: Primary tumor (rectum) and metastasis (liver) resection; 2009-2010: chemotherapy administration; 2011: metastatic relapse (liver) resection. CT images of metastatic sites are shown below each time point. **b**. Genotype matrix of somatic mutations identified in the malignant single-cells (gray=reference, blue=mutated, white=missing). Only non-silent mutations are shown (n=194), sorted according to their prevalence across the cell population. Gene names are displayed at the bottom of the map. Each row represents an individual cell. LRC1 is shown at the top of the matrix. Different colors highlight the tissue of origin - Purple: liver metastasis, Orange: liver relapse metastasis. **c**. Heatmap depicting genome-wide copy-number status (deletions: blue, amplifications: red) of tumor single-cells. Chromosomes ordered from 1 to 22, X and Y. Each row represents an individual cell, with the different colors highlighting the tissue of origin. Cell order is the same as in panel b.

We collected five tumor liver samples from the tumor and neighboring normal liver tissue at two different time points, from segment 7 in 2008 and segment 2 in 2011. All samples were snap-frozen in liquid nitrogen directly after surgical excision and stored at -80°C. Specifically, we obtained two tumor samples before treatment (Liver sample A [LA], Liver sample B [LB]), and three tumor samples after metastatic relapse resection (Liver Relapse sample A [LRA], LRB, and LRC). In addition, we collected one liver normal sample before treatment [NA].

### Tissue disaggregation and single-cell sorting

We minced each sample into 1-mm^3^ pieces with a scalpel and incubated them in Accutase (LINUS) for 1h at 37°C. We filtered the cell suspensions with a 70 μm cell strainer (Falcon, NY, USA) and assessed cell viability with Trypan Blue (Gibco, MA, USA). When the percentage of dead cells exceeded 30%, we performed a Ficoll-Paque density gradient centrifugation to discard dead cells and debris before sorting. We washed the cell pellets twice and re-suspended them in ice-cold phosphate-buffered saline (PBS). Then, we incubated them with antibodies against the epithelial cell adhesion molecule (EpCAM) (EBA1, FITC, 347197) and stem cell markers CD44 (APC, 559942), CD166 (PE, 559263), and LGR5 (BV 421, 562925) (all from BD Biosciences, NJ, USA). DRAQ5 (Thermo Fisher Scientific, MA, USA) and 7AAD (Invitrogen, MA, USA) dyes were additionally used to select nucleated cells and exclude non-viable ones. Individual tumor cells were gated and sorted into stem cell (EpCAM+/CD44+/CD166+/Lgr5-; EpCAM-/CD44-/CD166-/Lgr5+) and non-stem cell (EpCAM+/CD44-/CD166-/Lgr5-) phenotypes (Fig. S1). We carried out cell sorting using a FACSAria III (BD Biosciences, NJ, USA) flow cytometer and analyzed the resulting data using FACSDiva (BD Biosciences, NJ, USA) and FlowLogic software (Miltenyi Biotec, Germany).

### Single-cell whole-genome amplification

To obtain enough DNA for sequencing from each cell, we carried out single-cell whole genome amplification (scWGA) with the Ampli1 Kit (Menarini Silicon Biosystems, Italy). To avoid contamination, we worked under a biological safety cabinet, UV-irradiated all the plastic materials employed, and used a dedicated set of pipettes. In addition to the patient cells, we included a positive (10 ng/μl REPLIg human control kit, QIAGEN, Netherlands) and negative (DNase/RNase free water) control in the amplification process. We assessed the quality of the amplified DNA with the Ampli1 QC Kit. For positive samples for the four Ampli1 QC PCR fragments, we used the Ampli1 ReAmp/ds kit to increase the amount of total double-stranded DNA. After this, we removed the kit adaptors adding 5 μl of NE Buffer 4 10X (New England Biolabs, MA, USA), 1 μl of MseI 50U/μl (New England Biolabs, MA, USA), and 19 μl of nuclease-free water to every 25 μl of sample and incubated this mixture at 37ºC for 3 h, followed by 20 minutes at 65ºC for enzyme inactivation. After incubation, we purified the samples using 1.8X AMPure XP beads (Agencourt, Beckman Coulter, CA, USA). Finally, we measured the DNA yield with a Qubit 3.0 fluorometer (Thermo Fisher Scientific, MA, USA) and the amplicon size distribution with the D5000 ScreenTape assay in a 2200 TapeStation platform (Agilent Technologies, CA, USA).

### Bulk genomic DNA (gDNA) isolation

We isolated the gDNA from the normal bulk sample using the QIAamp DNA Mini kit (QIAGEN, Netherlands). Next, we estimated the DNA yield using the Qubit 3.0 fluorometer (Thermo Fisher Scientific, MA, USA) and the DNA integrity with the Genomic DNA ScreenTape Assay (Agilent Technologies, CA, USA).

### Next-generation sequencing

Single-cell and bulk libraries were constructed at the Spanish National Center for Genomic Analysis (CNAG; http://www.cnag.crg.eu) with the KAPA HyperPrep kit (Roche, Sweden). In total, 25 pre-treatment (18 LA + 7 LB) and 35 post-treatment (17 LRA + 17 LRB + 1 LRC) cells, plus one normal bulk sample (NA) were sequenced at ∼6X and ∼30X, respectively, on an Illumina NovaSeq 6000 (PE150) at CNAG.

### Single-cell variant calling

We aligned the single-cell and bulk sequencing reads to the Genome Reference Consortium Human Build 37 (GRCh37) using the MEM algorithm in the BWA software (*Website*, no date). Following a standardized best-practices pipeline (Van der Auwera *et al*., 2013), we filtered out reads displaying low mapping quality, performed local realignment around indels, and removed PCR duplicates. Under default settings, we identified somatic single nucleotide variants (SNVs) for each cell using SCcaller (Dong *et al*., 2017), a somatic variant caller specifically designed for scDNA-seq data. SNV calls were annotated using Annovar (v.20200608) (Wang, Li and Hakonarson, 2010). In parallel, we called single-cell copy-number variants (CNVs) with GINKGO (Garvin *et al*., 2015), using variable-length bins of around 500 kb; after binning, the data for each cell was normalized and segmented using default parameters. Afterward, we applied high-stringency thresholds to retain only high confidence calls, keeping mutations with a “True” somatic mutation tag and observed in a minimum of two single-cells. Then, we removed non-biallelic sites and sites missing in more than 50% of the sampled cells. In addition, genotypes within genomic regions affected by deletion events were converted to missing data.

### Mutational signature analysis

We ran sigProfilerExtractor (Ashiqul Islam *et al*., 2021) under default parameters to identify *de novo* SBS96 mutational signatures in the single cells and then assigned decomposed signatures to known ones COSMICv3 (Alexandrov *et al*., 2020).

### Phylogenetic analysis

We performed a Bayesian phylogenetic analysis using BEAST2 (2.6.4) (Bouckaert *et al*., 2019) under a relaxed clock model with exponential growth and with tip dates as days before present (with present-day being the last sampling date). Since our input consisted of a single-cell genotype matrix comprising variable genomic positions, we specified an SNV ascertainment bias correction by modifying the *constantPatterns* attribute in the input XML file. We ran a single MCMC chain during 500 million generations, recording samples every 1000 generations, and assessed convergence with Tracer v1.7.1 (Rambaut *et al*., 2018). After discarding the first 10% of the samples as burn-in, we obtained point estimates for the different parameters using posterior means. Then, we constructed a maximum clade credibility (MCC) tree using the median node heights. Finally, we mapped non-synonymous SNVs onto the MCC tree using the single-cell substitution model implemented in CellPhy (Kozlov *et al*., 2020).

### Demographic inference

We inferred the demographic history of the metastatic cells using Bayesian nonparametric phylodynamic reconstruction with preferential sampling (BNPR_PS). This approach tries to avoid the bias introduced by situations in which the sampling times depend on the effective population size (Karcher *et al*., 2017), as might be the case here. We used the MCC tree as input for the Phylodyn R package (https://github.com/mdkarcher/phylodyn).

### Selection analyses

We obtained the coding sequences for every cell using the dndscv R package (Martincorena *et al*., 2018) and concatenated them into a multiple sequence alignment. We then used the RELAX framework (Wertheim *et al*., 2015) to identify shifts in the non-synonymous/synonymous substitution rate ratio (dN/dS) along the cell phylogeny. Specifically, we tested whether differences in dN/dS between the branch leading to the most recent common ancestor of the relapsed cells and the remaining branches of the cell phylogeny could be attributed to changes in the intensity of selection. For this purpose, RELAX estimates a selection intensity parameter (*k*) and performs a likelihood ratio test to measure the significance of the results.

## Results

### The relapse population descends from a single lineage that survived treatment

Across all samples, we identified a total of 39,380 somatic SNVs, among which 194 were non-synonymous (**Fig. 1b**). We also found many CNVs across all malignant cells (**Fig. 1c**) with several marked differences in large-scale events between the pretreatment and relapsed cells (e.g., gains in chr 7, 11p, 20, and X). The phylogenetic analysis of the complete set of SNVs clearly showed three distinct and well-supported clades, strongly supporting a monoclonal origin of the relapse (**Fig. 2a**). Inside each clade, cells did not group according to their sampling location. Notably, 12 non-synonymous SNVs appear to be clonal (i.e., they are shared by all cells), including mutations in *APC, TP53* as well as several others affecting genes associated with increased tumor growth and poor prognosis in CRC and multiple cancer types, such as *IL23R* (Suzuki *et al*., *2012*), *PARP1* (Nosho *et al*., 2006), and *PSMA7* (Yang *et al*., 2013). Interestingly, the ancestral relapse lineage acquired a total of 516 mutations. Four of these mutations are non-synonymous, two of which occur in genes previously linked to CRC progression, *PKHD1* (Hu *et al*., 2016) and *CSF1R* (Lin *et al*., 2020). We estimated the age of the most recent common ancestor of all the metastatic cells sampled (mMRCA) to be 2.27 years, with a 95% highest posterior density (HPD) of 2.44-2.19 years. Interestingly, our results suggest that the relapse lineage started to diversify around 54 days (95% HPD: 22 - 112 days) before the surgical resection of the metastatic relapse.

**Figure 2.**
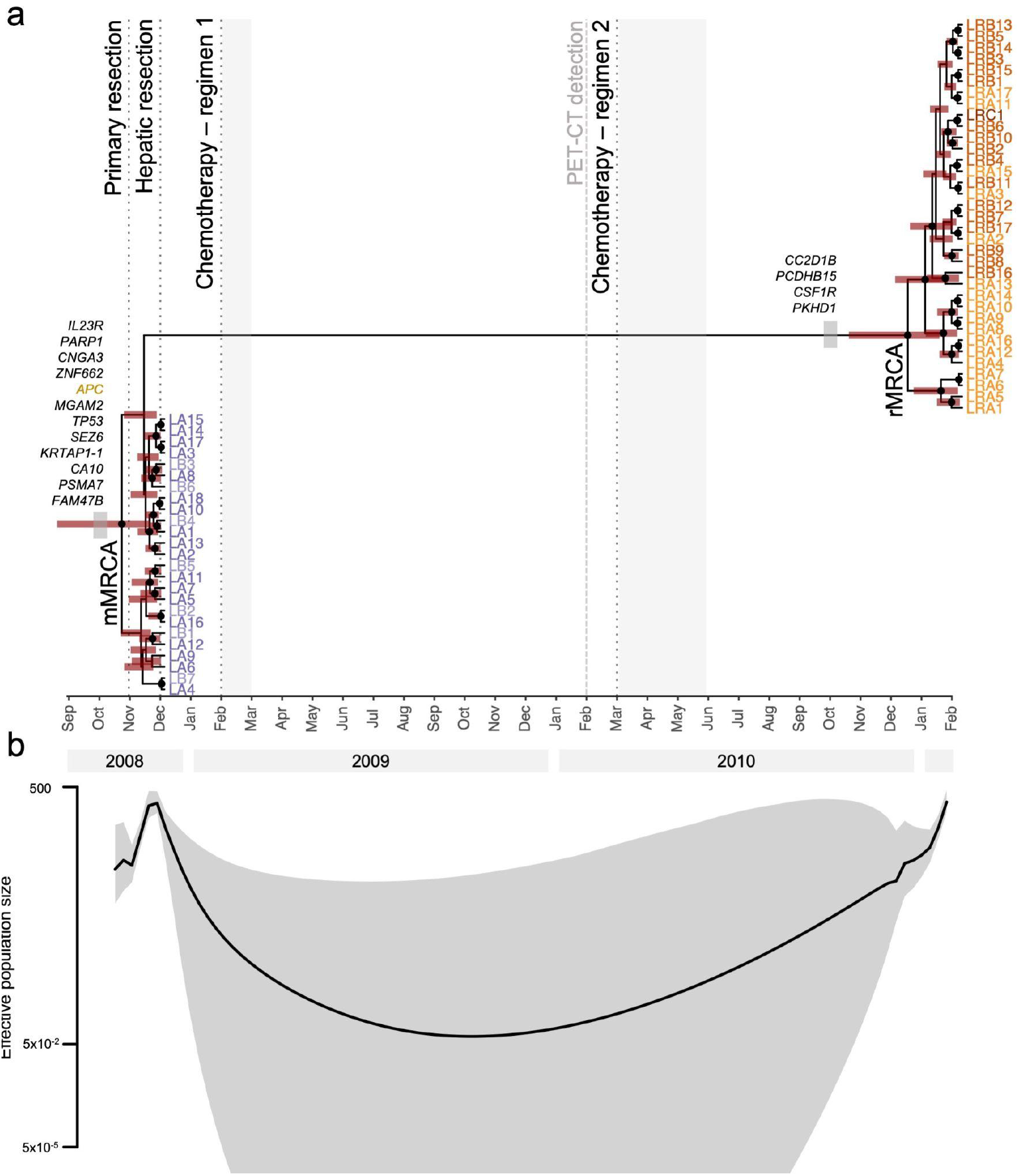
Phylogenetic and phylodynamic inference. **a**. Bayesian time-calibrated phylogeny. Tree nodes with posterior probability values > 0.9 are indicated with solid black circles. Red horizontal bars correspond to the 95% highest posterior density intervals of the age estimates. Solid gray horizontal bars depict the duration of each chemotherapy regimen. Single-cell IDs are shown at the tips of the tree. The x-axis is scaled in months. Non-silent mutations (missense in black and nonsense in gold) are displayed on the branches leading to the most recent common ancestor of the metastasis (mMRCA) and relapse (rMRCA) clades **b**. Changes in the effective population size of the metastatic population over time. The shaded area represents the 95% credible interval. The y-axis is in log scale.

### The relapse lineage evolved with fewer cells, slowly and under negative selection

We expect treatment to induce demographic changes in the tumor cell population. The Bayesian phylodynamic analysis (BNPR_PS) suggests that, following an initial expansion that led to the first metastasis, the tumor cell population underwent a sudden and sharp decline –coincident with the hepatic resection– reaching a flat stationary period that lasted until November 2009, after which the tumor cells began steadily expanding again, forming the relapse (**Fig. 2b**). Remarkably, these results agree with the imaging reports indicating an expansion of tumor cells already in February 2010. Moreover, the relaxed molecular clock analysis resulted in a global substitution rate of 4.31E-8 (median; per site per generation), falling close to previous estimates in CRC (Jones *et al*., 2008; Hu *et al*., 2019). However, the substitution rate does not seem to have been constant; we inferred substitution rate changes along the inferred phylogeny spanning over three orders of magnitude (1.8E-10 - 2.3E-07) (**Fig.3a**). Remarkably, the substitution rate was markedly low on the branch leading to the relapse population. Interestingly, the RELAX analysis points to this branch being under strong negative selection (dN/dS = 0.26) while the rest of the tree would be under slight positive selection (dN/dS = 1.27) (**Fig.3b**). However, this result was not statistically significant (k=1.17, LRT p-value = 0.999).

**Figure 3.**
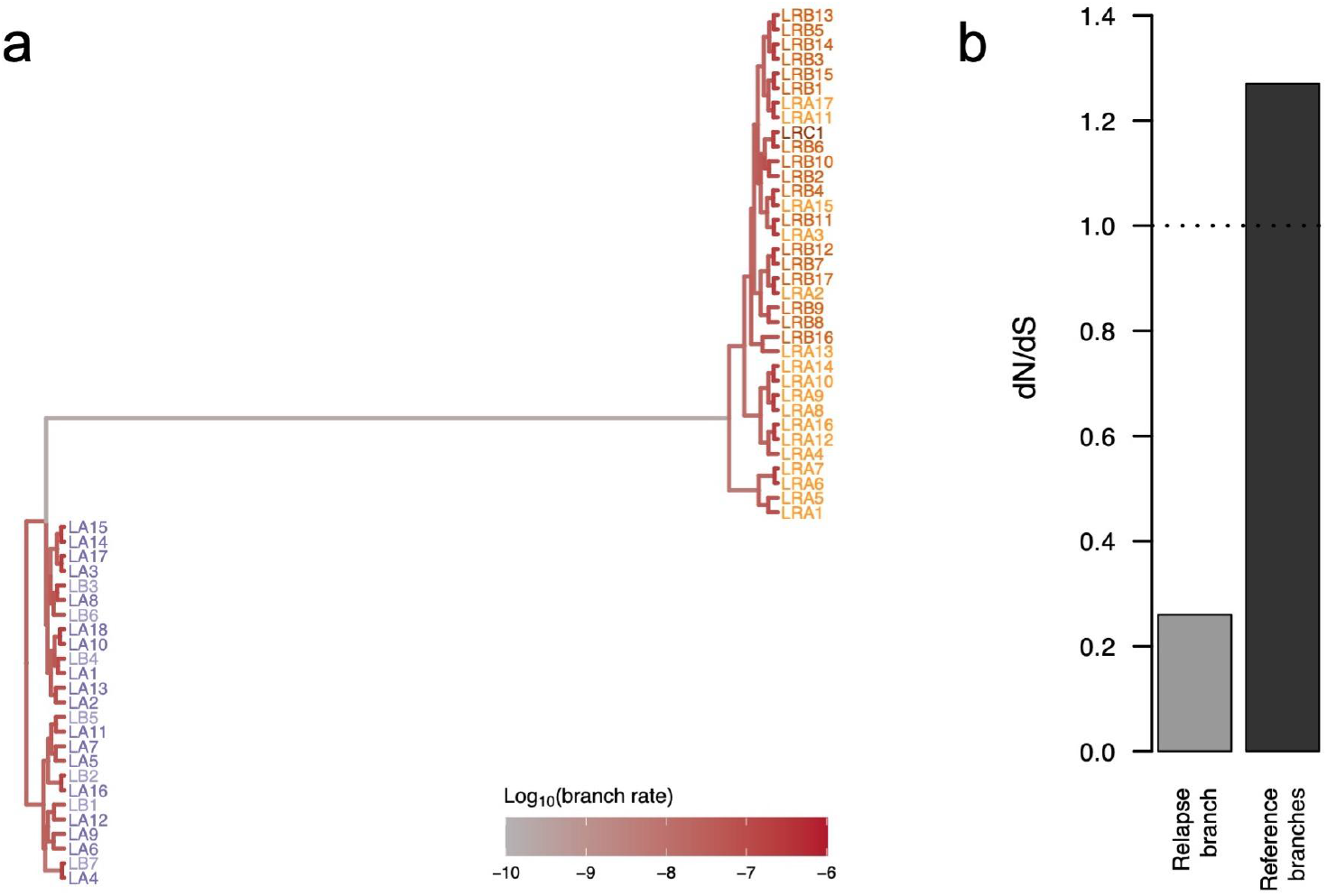
Branch rate variation during cancer evolution. **a**. Posterior substitution rate estimates per branch. Branch colors indicate the estimated substitution rate (low=gray, high=red). **b**. The bar plot depicts the dN/dS estimate obtained for the branch leading to relapse (gray) and the mean dN/dS estimate for the remaining branches of the tree (red). The dashed horizontal line highlights the neutral expectation of dN/dS = 1.

### Cells before and after relapse are subject to distinct mutational processes

We also explored the mutational signatures in the sampled cell populations to look for potential differences along the course of the disease. Of the four *de novo* mutational signatures that we identified (**Fig.4**, Fig. S2), signature 96-B was particularly active in the relapse cells, suggesting differences in the mutational processes between the first metastasis and the relapse lesion (**Fig.4a**). This signature, which is predominantly characterized by T>G mutations in a CTT trinucleotide context (**Fig.4b**), was further decomposed into COSMIC SBS signatures and found to be enriched in “clock-like” (i.e., aging) signatures SBS1 and SBS5, as well as signatures SBS35 and SBS17b (**Fig.4c**).

**Figure 4.**
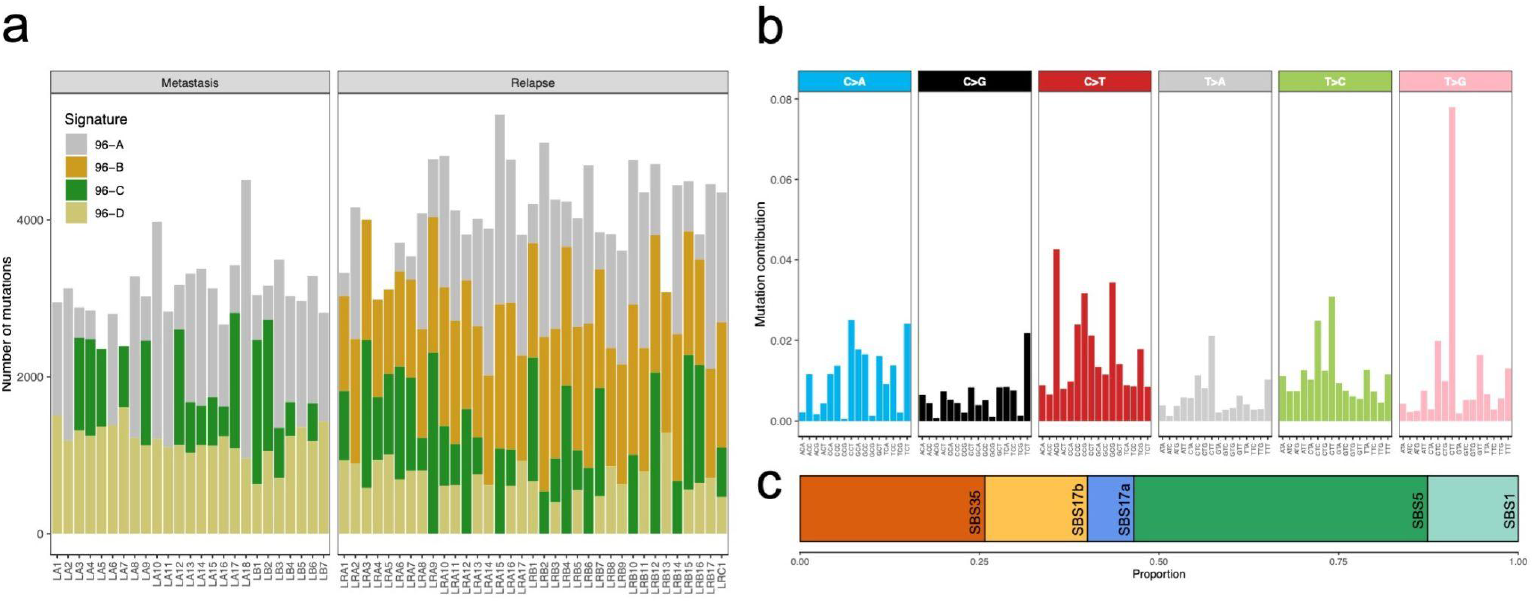
Mutational signatures across sampled single-cells. **a**. Barplot depicting the number of mutations contributing to the different signatures/processes across the different single cells. Cells sorted according to the tissue of origin with Cell IDs are shown at the bottom. Different colors reflect the identified mutational signatures. **b**. 96 trinucleotide substitution profiles of the 96-B mutational signature identified in the sampled relapse cells. **c**. COSMIC SBS deconvolution results for signature 96-B.

## Discussion

While primary CRC tumors have been extensively characterized from a genomic point of view (Cancer Genome Atlas Network, 2012), there have been far fewer analyses of metastatic and relapse CRC lesions, particularly with intervening systemic therapy in the course of the disease. Consequently, the time of onset and the molecular events underlying metastatic relapse have so far been difficult to characterize (Davis, Gao and Navin, 2017). Here, we used a unique dataset comprising 60 single-cell whole genomes derived from multiple samples, before and after treatment, from a liver metastasis of a CRC patient to explore the origins and evolution of the relapse in this individual.

In line with previous studies (Vogelstein and Kinzler, 2015), we observed an extensive genomic heterogeneity within the sampled lesions, including clonal mutations in genes often affected during the early stages of CRC development and relapse-specific mutations in genes previously associated with CRC progression and invasiveness. Moreover, our analyses suggest that the metastatic population underwent a strong bottleneck shortly after hepatic resection, declining further throughout the first chemotherapy regimen. Nevertheless, a single, resistant metastatic lineage seemed to expand again in late 2009. In addition, the fact that the inferred demographics, together with the PET-CT scan, indicate a tumor regrowth preceding the second regimen of chemotherapy suggests that the existence of a resistant subpopulation before treatment may have facilitated a more or less rapid relapse.

Our results suggest that a minor subclone survived treatment and likely started growing well before the clinical manifestation of the relapsing lesion in the liver. Detecting such minor clones can be very challenging unless continued monitoring for tumoral DNA, for example, through liquid biopsy, is implemented.

The substitution rate seems to have been considerably reduced in the ancestral metastatic lineage that gave rise to the relapse lesion. Such a slow pace of genetic change is likely the result of a strong drug-imposed selective pressure, which transiently decreased the genetic diversity available while imposing a severe evolutionary constraint on phenotypic variation.

Finally, we detected a unique mutational pattern affecting the cancer cells from the relapse lesion characterized by a substantial contribution of COSMIC signatures SBS35 and SBS17b. While signature SBS35 has been linked to platinum-based therapies, signature SBS17b shows a clear resemblance to the “5-FU” signature, recently identified in CRC patients undergoing 5-Fluorouracil treatment (Christensen *et al*., 2019). Notably, the noticeable contribution of these two mutational signatures to the overall mutational burden of the relapse cells appears to indicate a strong mutagenic effect associated with treatment (Christensen *et al*., 2019; Pich *et al*., 2019).

## Conclusions

In conclusion, our analyses illustrate how longitudinal single-cell whole-genome sequencing data can provide relevant insights into the evolutionary and molecular mechanisms promoting metastatic progression in CRC. Our results indicate that a minor resistant clone responsible for the relapse can be present long before clinical manifestation, thus highlighting the importance of longitudinal monitoring.

## Data Availability

We have deposited raw whole-genome single-cell sequencing data, together with a healthy bulk sample, at the Sequence Read Archive database under the accession code (to be provided).

## List of abbreviations

CNV: Copy number variant
COSMIC: Catalog of somatic mutations in cancer
gDNA: Genomic DNA
HPD: Highest posterior density
MCC: Maximum clade credibility
MCMC: Markov chain Monte Carlo
mCRC: Metastatic colorectal cancer
scDNA-seq: Single-cell DNA sequencing
scWGA: Single-cell whole genome amplification
SNV: Single nucleotide variant
XML: Extensible markup language

## Declarations

### Ethics approval and consent to participate

Tissue collection was performed at Charité - Universitätsmedizin Berlin and processed according to the Charité ethics vote EA1/187/16.

### Authors’ contributions

D.P. and S.M. conceived the study. D.P. and J.M.A. designed the analyses. C.S., N.R, D.G., J.P, and D.M obtained the tumor samples and patient information. S.P.L. and N.E.G. processed the samples, obtained the single-cells, and performed whole-genome amplification. J.M.A., L.T., and M.V. performed the analyses. D.G., N.K. and D.M. collected patient information and medical reports. All authors contributed to manuscript writing. All authors read and approved the final manuscript.

## Acknowledgments

We would like to thank all members of the phylogenomics lab for their comments on earlier versions of the manuscript. We also thank the Supercomputation Center of Galicia (CESGA) for providing all computational resources. We thank Dr. med. Bert Hildebrandt (Klinik für Gastroenterologie, Hämatologie und Internistische Onkologie GLG Werner Forßmann Klinikum Eberswalde) for clinical feedback.

## References

Alexandrov, L.B. et al. (2020) ‘The repertoire of mutational signatures in human cancer’, Nature, 578(7793), pp. 94–101.

Alves, J.M. et al. (2019) ‘Rapid evolution and biogeographic spread in a colorectal cancer’, Nature communications, 10(1), p. 5139.

Ashiqul Islam, S.M. et al. (2021) ‘Uncovering novel mutational signatures by de novo extraction with SigProfilerExtractor’, bioRxiv. doi:10.1101/2020.12.13.422570.

Bouckaert, R. et al. (2019) ‘BEAST 2.5: An advanced software platform for Bayesian evolutionary analysis’, PLoS computational biology, 15(4), p. e1006650.

Cancer Genome Atlas Network (2012) ‘Comprehensive molecular characterization of human colon and rectal cancer’, Nature, 487(7407), pp. 330–337.

Chen, H.-N. et al. (2022) ‘Genomic evolution and diverse models of systemic metastases in colorectal cancer’, Gut, 71(2), pp. 322–332.

Chen, J. et al. (2022) ‘Single cell DNA-seq depicts clonal evolution of multiple driver alterations in osimertinib resistant patients’, Annals of oncology: official journal of the European Society for Medical Oncology / ESMO [Preprint]. doi:10.1016/j.annonc.2022.01.004.

Christensen, S. et al. (2019) ‘5-Fluorouracil treatment induces characteristic T>G mutations in human cancer’, Nature communications, 10(1), pp. 1–11.

D’Angelica, M. et al. (2011) ‘Effect on outcome of recurrence patterns after hepatectomy for colorectal metastases’, Annals of surgical oncology, 18(4), pp. 1096–1103.

Davis, A., Gao, R. and Navin, N. (2017) ‘Tumor evolution: Linear, branching, neutral or punctuated?’, Biochimica et biophysica acta, 1867(2), p. 151.

Dong, X. et al. (2017) ‘Accurate identification of single-nucleotide variants in whole-genome-amplified single cells’, Nature methods, 14(5), pp. 491–493.

Garvin, T. et al. (2015) ‘Interactive analysis and assessment of single-cell copy-number variations’, Nature methods, 12(11), pp. 1058–1060.

van der Geest, L.G.M. et al. (2015) ‘Nationwide trends in incidence, treatment and survival of colorectal cancer patients with synchronous metastases’, Clinical &experimental metastasis, 32(5), pp. 457–465.

Hu, B. et al. (2016) ‘Abstract 3675: Loss of Pkhd1 promotes intestinal tumorigenesis in Apc mice’, Cancer research, 76(14 Supplement), pp. 3675–3675.

Hu, Z. et al. (2019a) ‘Quantitative evidence for early metastatic seeding in colorectal cancer’, Nature genetics, 51(7), pp. 1113–1122.

Imai, K. et al. (2016) ‘Early Recurrence After Hepatectomy for Colorectal Liver Metastases: What Optimal Definition and What Predictive Factors?’, The oncologist, 21(7), pp. 887–894.

Ishaque, N. et al. (2018) ‘Whole genome sequencing puts forward hypotheses on metastasis evolution and therapy in colorectal cancer’, Nature communications, 9(1), p. 4782.

Jones, S. et al. (2008) ‘Comparative lesion sequencing provides insights into tumor evolution’, Proceedings of the National Academy of Sciences of the United States of America, 105(11), pp. 4283–4288.

Karcher, M.D. et al. (2017) ‘phylodyn : an R package for phylodynamic simulation and inference’, Molecular Ecology Resources, pp. 96–100. doi:10.1111/1755-0998.12630.

Kim, C. et al. (2018) ‘Chemoresistance Evolution in Triple-Negative Breast Cancer Delineated by Single-Cell Sequencing’, Cell, 173(4), pp. 879–893.e13.

Kopetz, S. et al. (2009) ‘Improved survival in metastatic colorectal cancer is associated with adoption of hepatic resection and improved chemotherapy’, Journal of clinical oncology: official journal of the American Society of Clinical Oncology, 27(22), pp. 3677–3683.

Kozlov, A.M. et al. (2020) ‘CellPhy: accurate and fast probabilistic inference of single-cell phylogenies from scDNA-seq data’, bioRxiv. bioRxiv. doi:10.1101/2020.07.31.230292.

Leung, M.L. et al. (2017) ‘Single-cell DNA sequencing reveals a late-dissemination model in metastatic colorectal cancer’, Genome research, 27(8), pp. 1287–1299.

Levin, T.R. et al. (2018) ‘Effects of Organized Colorectal Cancer Screening on Cancer Incidence and Mortality in a Large Community-Based Population’, Gastroenterology, pp. 1383–1391.e5. doi:10.1053/j.gastro.2018.07.017.

Li H. (2013) ‘Aligning sequence reads, clone sequences and assembly contigs with BWA-MEM’, 1303:3997v1.

Lin, P.-C. et al. (2020) ‘Intratumor Heterogeneity of MYO18A and FBXW7 Variants Impact the Clinical Outcome of Stage III Colorectal Cancer’, Frontiers in oncology, 10, p. 588557.

Liu, D. et al. (2021) ‘Evolution of delayed resistance to immunotherapy in a melanoma responder’, Nature medicine, 27(6), pp. 985–992.

Martincorena, I. et al. (2018) ‘Universal Patterns of Selection in Cancer and Somatic Tissues’, Cell, 173(7), p. 1823.

Mitry, E. et al. (2008) ‘Adjuvant chemotherapy after potentially curative resection of metastases from colorectal cancer: a pooled analysis of two randomized trials’, Journal of clinical oncology: official journal of the American Society of Clinical Oncology, 26(30), pp. 4906–4911.

Nikbakht, H. et al. (2020) ‘Latency and interval therapy affect the evolution in metastatic colorectal cancer’, Scientific reports, 10(1), p. 581.

Nosho, K. et al. (2006) ‘Overexpression of poly(ADP-ribose) polymerase-1 (PARP-1) in the early stage of colorectal carcinogenesis’, European journal of cancer, 42(14), pp. 2374–2381.

Pich, O. et al. (2019) ‘The mutational footprints of cancer therapies’, Nature genetics, 51(12), pp. 1732–1740.

Rambaut, A. et al. (2018) ‘Posterior Summarization in Bayesian Phylogenetics Using Tracer 1.7’, Systematic Biology, pp. 901–904. doi:10.1093/sysbio/syy032.

Roh, V. et al. (2018) ‘Cellular Barcoding Identifies Clonal Substitution as a Hallmark of Local Recurrence in a Surgical Model of Head and Neck Squamous Cell Carcinoma’, Cell reports, 25(8), pp. 2208–2222.e7.

Sakamoto, H. et al. (2020) ‘The Evolutionary Origins of Recurrent Pancreatic Cancer’, Cancer discovery, 10(6), pp. 792–805.

Sentís, I. et al. (2020) ‘The evolution of relapse of adult T cell acute lymphoblastic leukemia’, Genome biology, 21(1), p. 284.

Shlush, L.I. et al. (2017) ‘Tracing the origins of relapse in acute myeloid leukaemia to stem cells’, Nature, 547(7661), pp. 104–108.

Sorbye, H. (2014) ‘Recurrence patterns after resection of liver metastases from colorectal cancer’, Recent results in cancer research. Fortschritte der Krebsforschung. Progres dans les recherches sur le cancer, 203, pp. 243–252.

Suzuki, H. et al. (2012) ‘IL-23 directly enhances the proliferative and invasive activities of colorectal carcinoma’, Oncology letters, 4(2), p. 199.

Van Cutsem, E. et al. (2016) ‘ESMO consensus guidelines for the management of patients with metastatic colorectal cancer’, Annals of oncology: official journal of the European Society for Medical Oncology / ESMO, 27(8), pp. 1386–1422.

Van der Auwera, G.A. et al. (2013) ‘From FastQ data to high confidence variant calls: the Genome Analysis Toolkit best practices pipeline’, Current protocols in bioinformatics / editoral board, Andreas D. Baxevanis … [et al.], 43, pp. 11.10.1–11.10.33.

Vogelstein, B. and Kinzler, K.W. (2015) ‘The Path to Cancer — Three Strikes and You’re Out’, New England Journal of Medicine, pp. 1895–1898. doi:10.1056/nejmp1508811.

Waanders, E. et al. (2020) ‘Mutational landscape and patterns of clonal evolution in relapsed pediatric acute lymphoblastic leukemia’, Blood cancer discovery, 1(1), pp. 96–111.

Wang, K., Li, M. and Hakonarson, H. (2010) ‘ANNOVAR: functional annotation of genetic variants from high-throughput sequencing data’, Nucleic acids research, 38(16), p. e164.

Wertheim, J.O. et al. (2015) ‘RELAX: Detecting Relaxed Selection in a Phylogenetic Framework’, Molecular Biology and Evolution, pp. 820–832. doi:10.1093/molbev/msu400.

Xi, Y. and Xu, P. (2021) ‘Global colorectal cancer burden in 2020 and projections to 2040’, Translational oncology, 14(10), p. 101174.

Yang, L. et al. (2013) ‘PSMA7 directly interacts with NOD1 and regulates its function’, Cellular physiology and biochemistry: international journal of experimental cellular physiology, biochemistry, and pharmacology, 31(6), pp. 952–959.

